# SARS-CoV-2 infection during the Omicron surge among patients receiving dialysis: the role of circulating receptor-binding domain antibodies and vaccine doses

**DOI:** 10.1101/2022.03.15.22272426

**Authors:** Maria E. Montez-Rath, Pablo Garcia, Jialin Han, LinaCel Cadden, Patti Hunsader, Curt Morgan, Russell Kerschmann, Paul Beyer, Mary Dittrich, Geoffrey A Block, Shuchi Anand, Julie Parsonnet, Glenn M Chertow

**Affiliations:** Department of Medicine (Nephrology), Stanford University; Ascend Clinical Laboratory; US Renal Care; Departments of Medicine (Infectious Diseases and Geographic Medicine), and Epidemiology and Population Health, Stanford University; Departments of Medicine (Nephrology), and Epidemiology and Population Health, Stanford University

## Abstract

**Background:** It is unclear whether a third dose of mRNA platform vaccines, or antibody response to prior infection or vaccination confer protection from the Omicron variant among patients receiving dialysis.

**Methods:** Monthly since February 2021, we tested plasma from 4,697 patients receiving dialysis for antibodies to the receptor-binding domain (RBD) of SARS-CoV-2. We assessed semiquantitative median IgG index values over time among patients vaccinated with at least one dose of the two mRNA vaccines. We ascertained documented COVID-19 diagnoses after December 25, 2021 and up to January 31, 2022. We estimated the relative risk for documented SARS-CoV-2 infection by vaccination status using a log-binomial model accounting for age, sex, and prior clinical COVID-19. Among patients with RBD IgG index value available during December 1-December 24, 2021, we also evaluated the association between the circulating RBD IgG titer and risk for Omicron variant SARS-CoV-2 infection.

**Results:** Of the 4,697 patients we followed with monthly RBD assays, 3576 are included in the main analysis cohort; among these, 852 (24%) were unvaccinated. Antibody response to third doses was robust (median peak index IgG value at assay limit of 150, equivalent to 3270 binding antibody units/mL). Between December 25-January 31, 2022, SARS-CoV-2 infection was documented 340 patients (7%), 115 (36%) of whom were hospitalized. The final doses of vaccines were given a median of 272 (25^th^, 75^th^ percentile, 245-303) days and 58 (25^th^, 75^th^ percentile, 51-95) days prior to infection for the 1-2 dose and 3 dose vaccine groups respectively. Relative risks for infection were higher among patients without vaccination (RR 2.1 [95%CI 1.6, 2.8]), and patients with 1-2 doses (RR 1.3 [95%CI 1.0, 1.8]), compared with patients with three doses of the mRNA vaccines. Relative risks for infection were higher among patients with RBD index values < 23 (506 BAU/mL), compared with RBD index value ≥ 23 (RR 2.4 [95%CI 1.9, 3.0]). The higher risk for infection among patients with RBD index values < 23 was present among patients who received three doses (RR 2.1 [95%CI 1.3, 3.4]).

**Conclusions:** Among patients receiving hemodialysis, patients unvaccinated, without a third mRNA vaccine dose, or those lacking robust circulating antibody response are at higher risk for Omicron variant infection. Low circulating antibodies could identify the subgroup needing intensified surveillance, prophylaxis or treatment in this patient population.

## INTRODUCTION

Antibody response to COVID-19 vaccination after the primary series is diminished in up to 15% of patients receiving dialysis^1-4^. Among patients with an initial response, circulating antibody levels often wane^5-7^. Prior to the emergence of the highly transmissible Omicron (B.1.1.529) variant, we and others showed that low circulating antibody levels were linked with a greater than 10-fold increased risk for breakthrough infections^5,8^. In patients receiving dialysis, infection with SARS-CoV-2, even post-vaccination, often results in hospitalization^9,10^ and carries the additional risk for in-facility transmission^11^.

To date, only half of patients on dialysis have agreed to a third (booster) dose of the mRNA platform vaccines^12^. Although a third dose generates an antibody response in nearly all patients receiving dialysis^13,14^, the persistence of the response is unknown. Preliminary data on the clinical effectiveness of a third dose against the SARS-CoV-2 Omicron variant in this population are mixed^15,16^. Moreover, since the Omicron variant receptor binding domain (RBD) differs substantially from that of the progenitor (Wuhan) virus, post-vaccination or post-infection circulating antibody levels to the RBD may offer limited to no protection against infection.

In a prospective cohort of 4,697 patients receiving dialysis throughout the U.S. in whom we have tracked monthly SARS-CoV-2 antibody response since February 1, 2021, we evaluated the longitudinal circulating RBD antibody response among patients with one or two versus three doses of mRNA vaccines as of December 2021. We also evaluated the effectiveness of three mRNA vaccine doses, and determined the relations between circulating antibody response and subsequent Omicron breakthrough infection from December 25, 2021 to January 31, 2022, the period during which the SARS-CoV-2 Omicron variant was the dominant variant in the U.S.

## METHODS

Starting in February 2021, in partnership with a central laboratory (Ascend Clinical), we tested monthly remainder plasma samples from a cohort of patients receiving dialysis at U.S. Renal Care for RBD antibody. U.S. Renal Care is a dialysis network with more than 350 facilities nationwide. We have previously described sample size estimation and methods in detail. We used electronic health records to ascertain patient characteristics, vaccination status, and SARS-CoV-2 diagnosis. The study received ethics approval from Stanford University. Stanford University investigators received anonymized data, and the Institutional Review Board waived the requirement for consent.

### Patient population

We included patients who were unvaccinated and vaccinated with one, two, or three doses of one of two available mRNA vaccines, as reported in the electronic health record (see **STable 1** for distribution of vaccine combinations). We excluded patients who had received other vaccines due to limited numbers. We assigned type of vaccination by the first dose vaccine type.

### Documented SARS-CoV-2 infection

We ascertained SARS-CoV-2 infection using the U.S. Renal Care electronic health record of a documented SARS-CoV-2 infection^5^. We also extracted data on hospitalizations in the seven days before or the 14 days after the diagnosis date. For the purposes of evaluating infection risk during the period when the Omicron variant (BA1.1, B.1.1.529, BA.2) was dominant in the U.S., we evaluated SARS-CoV-2 infection diagnoses from December 25, 2021 through January 31, 2022. Centers for Disease Control and Prevention (CDC) data delineate that the Omicron variant was detected in 74% of U.S. infections by the last week of December, and increased rapidly thereafter^12^.

### Laboratory Testing for RBD Antibodies

We tested remainder samples using the Siemens total RBD Ig assay, which measures IgG and IgM antibodies. The assay is reported by the manufacturer to have 100% sensitivity and 99.8% specificity if performed 14 or more days after a positive result on a reverse transcriptase polymerase chain reaction (RT-PCR) test^17^, and has been independently validated, with similar performance characteristics^18^. On a monthly basis starting in February 2021, we used this assay to test remainder samples from patients in whom antibodies had not been detected in the prior month. After a positive total RBD Ig result, the positive sample and all subsequent monthly samples were tested using a semiquantitative Siemens RBD IgG assay (Atellica IM sCOVG assay)^17^. This assay is a 2-step sandwich indirect chemiluminescent assay with manufacturer-reported sensitivity of 95.6% (95% CI, 92.2% to 97.8%) and specificity of 99.9% (CI, 99.6% to 99.9%) for tests performed 21 or more days after a positive RT-PCR result. An index value of 1.0 corresponds to 21.8 binding antibody units (BAU) per milliliter according to the recently established World Health Organization (WHO) international standard^19^. An index value of 1.0 or greater (≥21.8 BAU/mL) is considered reactive on this assay, and an index value of 150 (3270 BAU/mL) is the upper limit of quantification.

### Statistical Analysis

Two of the authors (M.E.M. and J.H.) led the statistical analyses. The main analysis includes all patients alive and on hemodialysis at one of the participating U.S. Renal Care facilities as of December 24, 2021. For determining risk of SARS-CoV-2 Omicron variant infection by circulating antibody titer, we restricted further to patients with an RBD antibody titer available during December 1-December 24, 2021 (**Figure 1**).

**Figure 1.**
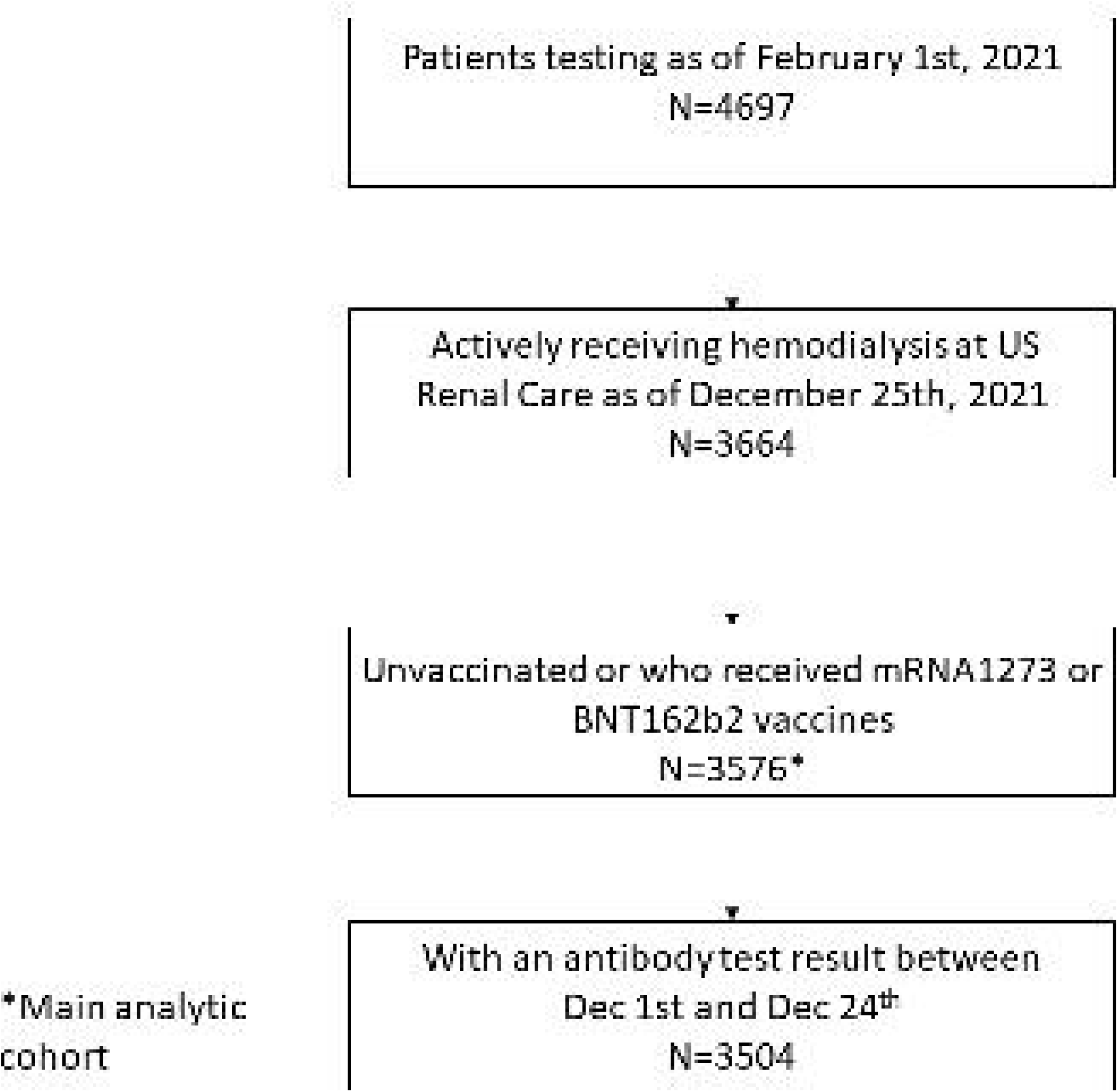
Flow chart of main analysis cohort creation.

We described the cohort by vaccination status—unvaccinated, completed initial series, and completed booster—using proportions, means and SDs, or medians and interquartile ranges, as applicable. Next, among patients with at least one dose of the mRNA1273 or BNT162b2, we described the antibody response over time by type of vaccine (defined by the vaccine type received as the first dose) and vaccination status. We estimated age- and sex-adjusted medians and 95% CIs for RBD IgG index values using quantile regression with robust SEs to account for multiple observations per patient, as implemented in the Stata qreg and margins commands. In this longitudinal data analysis, model parameters have a population-average interpretation. We used quantile regression and, in particular, the median to describe the data because it is invariant to data truncation and estimable in all of the analyses presented. Because patients receiving dialysis are tested monthly on or around the same date each month, we reported data using discrete post-vaccination 30-day time windows. To describe the full distribution of index values in each period by vaccination type and status, we produced boxplots of the RBD IgG index values.

Finally, we estimated the relative risk for documented SARS-CoV-2 infection by vaccination status using a log-binomial model accounting for age, sex, and prior documented SARS-CoV-2 infection. We followed similar procedures to evaluate the association between the circulating RBD IgG titer and risk for Omicron variant SARS-CoV-2 infection using the sub-cohort of patients with an RBD antibody titer available during December 1-December 24, 2021. We *a priori* selected circulating RBD threshold as < 23 versus ≥ 23 (506 BAU/mL), on the basis of our^1^ and others’^20^ prior data demonstrating higher risk for breakthrough infection post-vaccination at this thresholds.

We assumed statistical significance at a 2-sided α level of 0.05. Statistical analyses were conducted using SAS, version 9.4 (SAS Institute), or Stata/MP 17 (StataCorp).

## RESULTS

Of the 4,697 patients we followed as of February 1, 2021, we included 3,576 in the analytic cohort (**Figure 1**). Among these, 852 (24%) were unvaccinated (**Table 1**). Compared with patients who had received 1-2 doses of mRNA vaccination, the unvaccinated subgroup was younger, and more likely to be Non-Hispanic Black, to not have diabetes, and to reside in the South. Twenty five percent of the cohort received a third dose; this subgroup was older, and more likely to be Hispanic, to have diabetes, and to reside in the West, compared with patients receiving 1-2 doses.

**Table 1.**
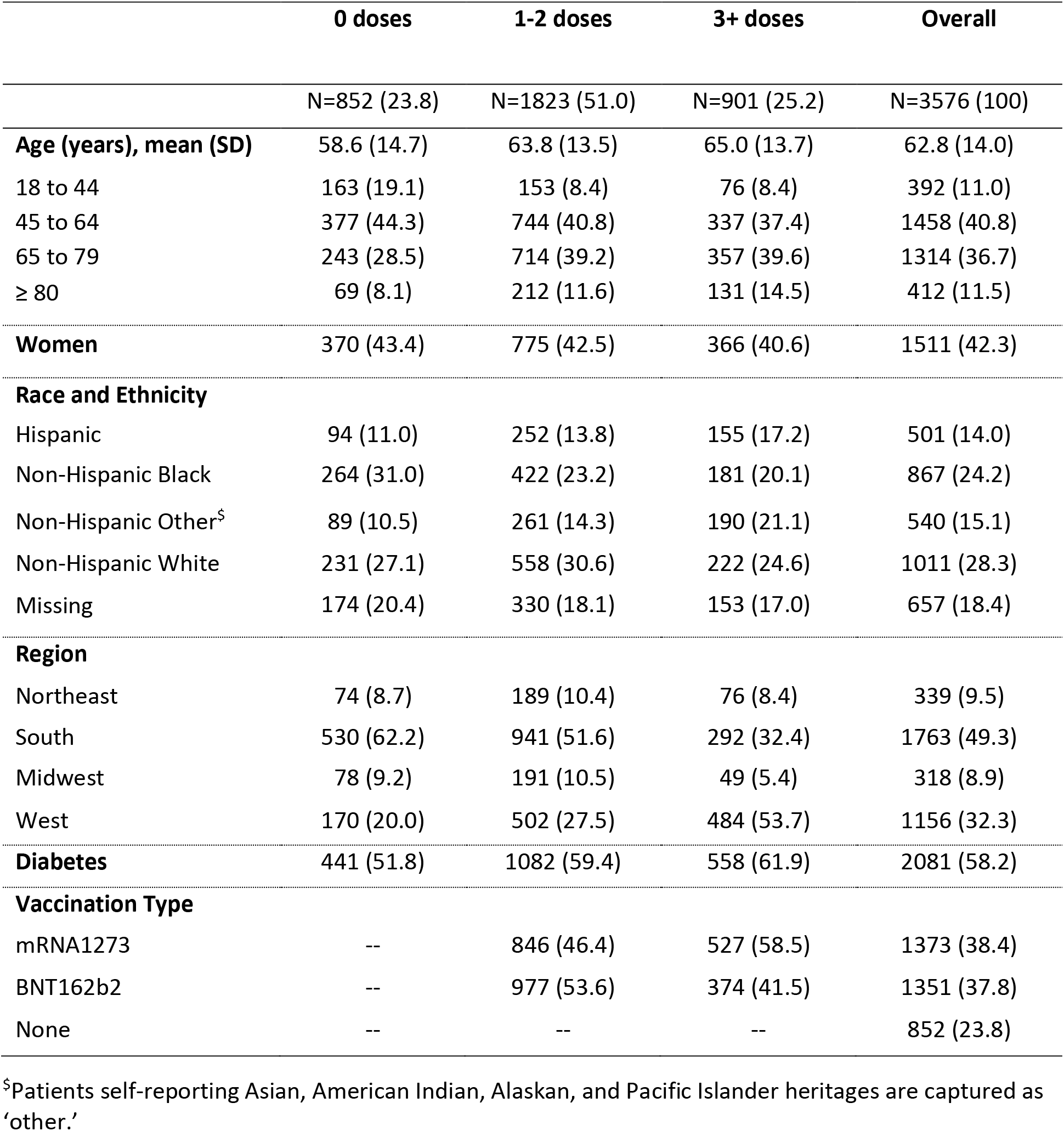
Characteristics of patients followed with monthly RBD antibody tests alive and on dialysis by vaccination status on Dec 24^th^ 2021.

Among the subgroup with RBD IgG index values available in December 2021, 1773 (51%) had RBD IgG index values < 23 (**STable 2**). The group with index value < 23 was younger, and more likely to be unvaccinated, to be Non-Hispanic white, and to not have diabetes.

### Antibody response to third vaccine dose

Prior to the third dose, median RBD IgG index values peaked between day 31-60 since vaccination and started to decrease by day 61-90, reaching a plateau during day 121-270 (**Figure 2A** and **2B**; **STable 3** for numbers of patients included in each time window; **SFigure 1A** and **1B** for box plots depicting the entire distribution of the data). Patients who received the mRNA1273 vaccine had higher median values throughout the follow up compared with patients who received the BNT162b2 vaccine.). After day 270, we observed an increase in the median index RBD values for patients with a third dose of the mRNA vaccines, concomitant with the timing of the third dose (median time to third dose 250 [25^th^, 75^th^ percentile 231, 280] days since first dose). Median peak RBD IgG index value response to both vaccines after third dose reached the assay limit (index value 150, 3270 BAU/mL).

**Figure 2.**
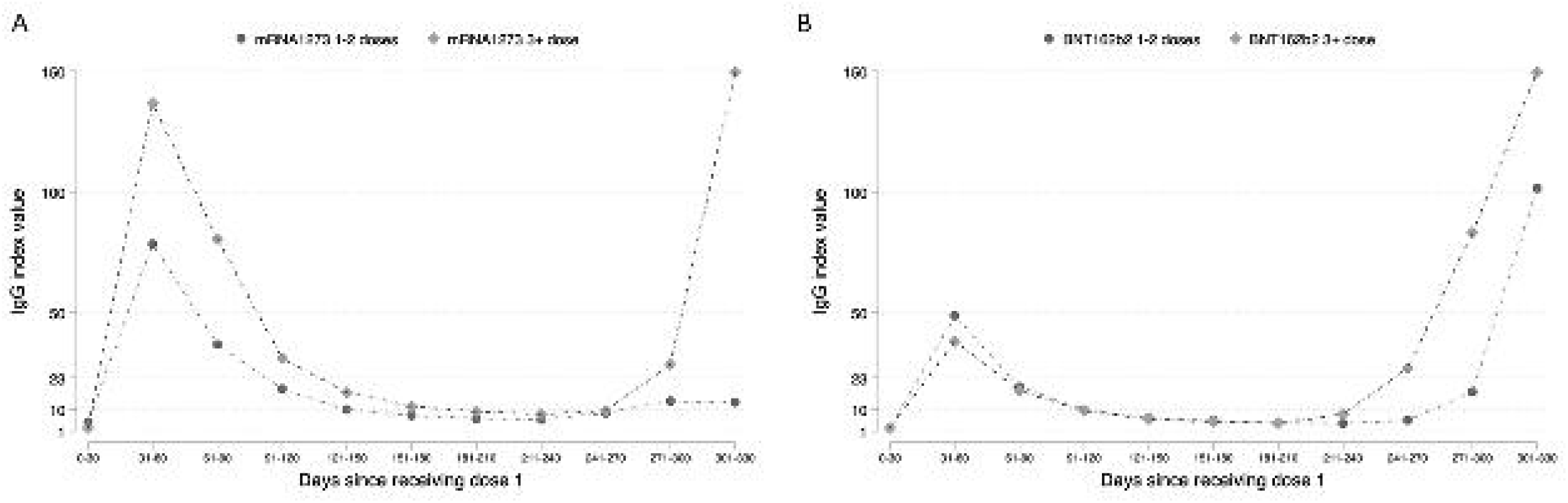
Age and Sex adjusted median antibody levels between Feb 1^st^ and Dec 24^th^, 2021, plotted from 1^st^ dose if vaccinated or from Feb 1^st^, 2021, if unvaccinated, measured in 30-day periods A. mRNA1273 B. BNT162b2.

Patients with 1-2 doses of mRNA1273 and BNT162b2 and also had an increase in median index values after day 270 window post-vaccination, with a greater increase in median index values observed among patients receiving 1-2 doses BNT162b2. We hypothesized that this increase may be related to SARS-CoV-2 breakthrough infection prior to December 25, 2021. A total of 28 patients had documented SARS-CoV-2 infection after day 270 and prior to December 25, 2021; of these 20 (71%) were in the 1-2 dose group.

### Risks for Omicron variant infection

A total of 340 (7%) patients had documented COVID-19 between December 25-January 31, 2022, and 115 (36%) of these were hospitalized in the one week prior to or two weeks after their infection diagnosis. The final doses of vaccines were given a median of 272 (25^th^, 75^th^ percentile 245, 303) days and 58 (25^th^, 75^th^ percentile 50, 95) days prior to infection for the 1-2 dose and 3 dose vaccine groups respectively. Relative risks for infection were higher among patients without vaccination, and patients with 1-2 doses compared with patients with three doses of the mRNA vaccines, after accounting for age, sex, and prior documented COVID-19 (**Table 2**).

**Table 2.** Relative risk of getting breakthrough infection by vaccine status (vs 3+ doses) and by RBD index value overall and by vaccination status.

|  | Unadjusted | Adjusted |  |
| --- | --- | --- | --- |
|  |  | Age and sex | Age, sex and prior documented SARS-CoV-2 infection |
| Vaccine doses |  |  |  |
| 0 | 2.0 (1.5, 2.7) | 2.0 (1.5, 2.7) | 2.1 (1.6, 2.8) |
| 1-2 | 1.3 (1.0, 1.8) | 1.3 (1.0, 1.8) | 1.3 (1.0, 1.8) |
| 3+ | Ref | Ref | Ref |
| RBD Status |  |  |  |
| <23 Vs ≥ 23 | 2.4 (1.9, 3.0) | 2.4 (1.9, 3.0) | 2.4 (1.9, 3.0) |
| Vaccine doses and RBD status<sup>§</sup> |  |  |  |
| 0 doses IgG < 23 | 3.1 (2.2,4.5) | 3.2 (2.2,4.6) | 3.2 (2.2,4.5) |
| 0 doses IgG ≥ 23 | 1.2 (0.7,2.2) | 1.3 (0.7,2.3) | 1.4 (0.8,2.4) |
| 1-2 doses IgG < 23 | 2.2 (1.6,3.2) | 2.3 (1.6,3.3) | 2.2 (1.6,3.2) |
| 1-2 doses IgG ≥ 23 | 1.1 (0.7,1.6) | 1.1 (0.7,1.6) | 1.1 (0.7,1.7) |
| 3 plus doses IgG < 23 | 2.1 (1.3,3.4) | 2.1 (1.3,3.5) | 2.1 (1.3,3.4) |
| 3 plus doses IgG ≥ 23 | Ref | Ref | Ref |

<sup>§</sup>Interaction p-value <0.001

Among the 3504 patients with an RBD IgG index value available during December 1-December 24, 2021, 339 (10%) patients had documented SARS-CoV-2 infection between December 25-January 31, 2022 with a median time between index values and infection of 37 (25^th^, 75^th^ percentile, 31-46) days. Relative risks for infection were higher among patients with RBD IgG index values < 23 (versus RBD IgG ≥ 23). In evaluating the interaction between RBD IgG and vaccination (**Table 2**), patients with three doses of vaccine and RBD IgG index values < 23 had lower risk for infection (adjusted RR 2.1 [95% CI 1.3, 3.4]) versus unvaccinated patients with RBD IgG index values < 23 (adjusted RR 3.2 [2.2, 4.5]), compared with patients with three doses of vaccine and RBD IgG ≥ 23. There were larger differences in relative risks by antibody response, regardless of number of vaccine doses (**Table 2**).

## DISCUSSION

In this nationwide cohort of patients receiving dialysis, among whom more than 75% had received at least one dose of the mRNA platform vaccines, 7% experienced documented SARS-CoV-2 infection during the Omicron variant surge period in the U.S. Unvaccinated patients, patients without a third vaccine dose, and those lacking robust circulating antibody response were at higher risk for infection. Even among patients receiving three or more doses of vaccine, RBD index values < 23 (< 508 BAU/mL) were associated with nearly 2-fold higher risk for breakthrough infection. Although increasing number of vaccine doses reduced risks for Omicron infection, the antibody response demonstrated a strong, independent association with infection risk, indicating its potential applicability for risk stratification in this patient population.

Our data are consistent with a study including 1121 patients receiving hemodialysis in the U.K. who were also undergoing weekly screening by rtPCR testing^14^. In this study, 12.9% of patients receiving hemodialysis experienced Omicron SARS-CoV-2 infection (symptomatic and asymptomatic) between December 1, 2021 and January 16, 2022. Compared to unvaccinated patients, patients with three, but not two, ChAdOx1 or BNT162b2 vaccine doses experienced a reduced likelihood of Omicron infection (HR 0.50 [95% CI 0.29-0.92])^15^. These findings are consistent with general population data from the U.K. Health Security Agency, in which the investigators reported suboptimal vaccine effectiveness against symptomatic Omicron variant SARS-CoV-2 among persons receiving two vaccine doses^21^. Carr et al. evaluated neutralizing antibody response against Omicron, compared with Delta, SARS-CoV-2 variants, among patients receiving dialysis^16^. Median neutralizing antibody titers against Omicron among patients with two doses of BNT162b2 were below assay range at day 158 after two doses, and rose significantly to detectable ranges at day 27 after three doses. Concordant with our findings, these studies suggest that three doses of the current formulation of the mRNA vaccines provide enhanced protection from Omicron variant SARS-CoV-2 infection in the short-term post third dose.

Continued illness and hospitalization of patients receiving dialysis despite vaccination, however, raises questions about durable effectiveness of the vaccines in this high-risk population, the correct vaccination dosing schedule, and the need for follow-up antibody testing. In the Carr et al. study, 25% of patients receiving dialysis did not mount neutralizing antibody titers against the Omicron variant in the immediate period post third dose^16^. Moreover, the duration of detectable neutralizing antibody titers and/or real-world protection against SARS-CoV-2 post third dose is unknown. Compared with solid organ transplant recipients or patients receiving cytotoxic or B-cell depleting chemotherapy in whom the CDC currently recommends a fourth dose^22^, patients receiving dialysis had higher rates of seroconversion after the initial vaccination series and after the third dose. Nonetheless, patients receiving dialysis experience higher rates of infection-related complications than the general population^9^. Measurement of circulating anti-SARS-CoV-2 antibody levels may help identify the subgroup of patients receiving dialysis that remains at higher risk post-vaccination, and may benefit from pre-exposure prophylaxis with monoclonal antibodies, for example, since the patient population on dialysis as a whole is not currently included in the moderate to severely immunocompromised group warranting prophylaxis^23^. Following precedence of hepatitis B vaccination, serial measurements of circulating antibody levels are feasible in patients receiving dialysis, and could inform timing for offering additional doses, monoclonal antibody prophylaxis or anti-viral treatment.

The strengths of our analysis include a diverse cohort (by age, sex, self-reported race/ethnicity, geography, socioeconomic status, and dialytic modality), and the use of a single, highly sensitive and specific assay over time. We are able to ascertain circulating antibody titers proximal to infection and re-evaluate previously suggested RBD antibody level thresholds in the context of a new variant.

We could not evaluate risks for Omicron variant COVID-19 among patients receiving Ad26.COV2.S vaccine as the initial series due to small numbers of patients receiving this formulation at U.S. Renal Care facilities. Since we are measuring RBD antibody levels, which respond both to infection and to vaccination, we cannot identify patients with asymptomatic SARS-CoV-2 infection who may have added immunity beyond that induced through vaccination.

In summary, we demonstrated a meaningful but incomplete degree of protection against infection with the SARS-CoV-2 Omicron variant following a third (booster) dose of mRNA platform vaccines. The risk for infection after third vaccine dose is dependent on the antibody response. Ongoing efforts to determine the optimal schedule of SARS-CoV-2 infection surveillance and the utility and timing of antibody testing in patients receiving dialysis should yield benefits to patients receiving dialysis and the public health at large.

## Data Availability

Data set: Available upon investigators' review of request, with age categories further collapsed to ensure anonymization. The analytical data set may be subject to a data sharing agreement with U.S. Renal Care.

## References

1. Anand S, Montez-Rath M, Han J, et al. Antibody Response to COVID-19 Vaccination in Patients Receiving Dialysis. J Am Soc Nephrol. 2021.

2. Garcia P, Anand S, Han J, et al. COVID-19 Vaccine Type and Humoral Immune Response in Patients Receiving Dialysis. J Am Soc Nephrol. 2021.

3. Grupper A, Sharon N, Finn T, et al. Humoral Response to the Pfizer BNT162b2 Vaccine in Patients Undergoing Maintenance Hemodialysis. Clin J Am Soc Nephrol. 2021.

4. Hasmann S, Paal M, Fuessl L, Fischereder M, Schonermarck U. Humoral immunity to SARS-CoV-2 vaccination in haemodialysis patients: (Response to: Humoral and cellular immunity to SARS-CoV-2 vaccination in renal transplant versus dialysis patients: A prospective, multicenter observational study using mRNA-1273 or BNT162b2 mRNA vaccine.). Lancet Reg Health Eur. 2021;10:100237.

5. Anand S, Montez-Rath ME, Han J, et al. SARS-CoV-2 Vaccine Antibody Response and Breakthrough Infection in Patients Receiving Dialysis. Ann Intern Med. 2021.

6. Berar-Yanay N, Freiman S, Shapira M, et al. Waning Humoral Response 3 to 6 Months after Vaccination with the SARS-COV-2 BNT162b2 mRNA Vaccine in Dialysis Patients. J Clin Med. 2021;11(1).

7. Hsu CM, Weiner DE, Manley HJ, et al. Seroresponse to SARS-CoV-2 Vaccines among Maintenance Dialysis Patients over 6 Months. Clin J Am Soc Nephrol. 2022;17(3):403–413.

8. Boudhabhay I, Serris A, Servais A, et al. COVID-19 outbreak in vaccinated patients from a hemodialysis unit: antibody titers as a marker of protection from infection. Nephrol Dial Transplant. 2022.

9. Salerno S, Messana JM, Gremel GW, et al. COVID-19 Risk Factors and Mortality Outcomes Among Medicare Patients Receiving Long-term Dialysis. JAMA Netw Open. 2021;4(11):e2135379.

10. Medicare Cf, Services M. Preliminary Medicare COVID-19 data snapshot. In:2021.

11. Corbett RW, Blakey S, Nitsch D, et al. Epidemiology of COVID-19 in an Urban Dialysis Center. J Am Soc Nephrol. 2020;31(8):1815–1823.

12. Centers for Disease Control and Prevention. COVID Data Tracker. 2022; https://covid.cdc.gov/covid-data-tracker/. Accessed March 7, 2022.

13. Garcia PH, J. Montez-Rath, Maria. Sun, Sumi. Shang, Tiffany. Parsonnet, Julie. Chertow, Glenn. Anand, Shuchi. Abra, Graham. Schiller, Brigitte. SARS-COV-2 booster vaccine response among patients receiving dialysis. Clinical Journal of the American Society of Nephrology. 2022.

14. Ducloux D, Colladant M, Chabannes M, Yannaraki M, Courivaud C. Humoral response after 3 doses of the BNT162b2 mRNA COVID-19 vaccine in patients on hemodialysis. Kidney Int. 2021;100(3):702–704.

15. Spensley K, Gleeson S, Martin P, et al. Comparison of vaccine effectiveness against the Omicron (B.1.1.529) variant in patients receiving haemodialysis. medRxiv. 2022.

16. Carr EJ, Wu M, Harvey R, et al. Omicron neutralising antibodies after COVID-19 vaccination in haemodialysis patients. Lancet. 2022.

17. U.S. Food & Drug Administration. EUA Authorized Serology Test Performance. 2020.

18. Schnurra C, Reiners N, Biemann R, Kaiser T, Trawinski H, Jassoy C. Comparison of the diagnostic sensitivity of SARS-CoV-2 nucleoprotein and glycoprotein-based antibody tests. J Clin Virol. 2020;129:104544.

19. Kristiansen PA, Page M, Bernasconi V, et al. WHO International Standard for anti-SARS-CoV-2 immunoglobulin. Lancet. 2021;397(10282):1347–1348.

20. Feng S, Phillips DJ, White T, et al. Correlates of protection against symptomatic and asymptomatic SARS-CoV-2 infection. Nat Med. 2021;27(11):2032–2040.

21. Andrews N, Stowe J, Kirsebom F, et al. Covid-19 Vaccine Effectiveness against the Omicron (B.1.1.529) Variant. New England Journal of Medicine. 2022.

22. Centers for Disease Control and Prevention. COVID-19 Vaccines for Moderately or Severely Immunocompromised People. https://www.cdcgov/coronavirus/2019-ncov/vaccines/recommendations/immunohtml. 2022;Last accessed March 14, 2022.

23. Food and Drug Administration. Frequently Asked Questions on the Emergency Use Authorization for Evusheld (tixagevimab co-packaged with cilgavimab) for Pre-exposure Prophylaxis (PrEP) of COVID-19. https://www.fdagov/media/154703/download. 2022;Last accessed March 14, 2022.

