## Supplement Table 1 and Table 2 for "SARS-CoV-2 infection during the Omicron surge among patients receiving dialysis: the role of circulating receptor-binding domain antibodies and vaccine doses"

**STable 1. Observed vaccine type combinations among the main cohort patients (sorted in decreasing order of observed percent).**

| Vaccine type combination | Count | Percent |
|---|---|---|
| U | 852 | 23.83 |
| BB | 728 | 20.36 |
| MM | 657 | 18.37 |
| MMM | 449 | 12.56 |
| BBB | 344 | 9.62 |
| B | 245 | 6.85 |
| M | 169 | 4.73 |
| MMB | 59 | 1.65 |
| MB | 20 | 0.56 |
| BBM | 20 | 0.56 |
| MBB | 7 | 0.20 |
| MMMM | 6 | 0.17 |
| MBM | 5 | 0.14 |
| BBBB | 5 | 0.14 |
| BM | 4 | 0.11 |
| MBBB | 1 | 0.03 |
| BMM | 1 | 0.03 |
| BMMB | 1 | 0.03 |
| BMB | 1 | 0.03 |
| BBMB | 1 | 0.03 |
| BBBM | 1 | 0.03 |

U: Unvaccinated; M: mRNA1273; B: BNT162b2.

**STable 2. Characteristics of patients with a RBD antibody test performed between Dec 1st and Dec 24th 2021 alive and on dialysis by IgG level on Dec 24th 2021.**

| | **IgG index <23** | **IgG index >= 23** | **Overall** |
|---|---|---|---|
| | N=1773* (50.6) | N=1731 (49.4) | N=3504 (100) |
| **Age (years), mean (SD)** | 61.6 (14.1) | 64.0 (13.9) | 62.8 (14.0) |
| 18 to 44 | 214 (12.1) | 169 (9.8) | 383 (10.9) |
| 45 to 64 | 783 (44.2) | 654 (37.8) | 1437 (41.0) |
| 65 to 79 | 598 (33.7) | 681 (39.3) | 1279 (36.5) |
| ≥ 80 | 178 (10.0) | 227 (13.1) | 405 (11.6) |
| **Women** | 758 (42.8) | 722 (41.7) | 1480 (42.2) |
| **Race and Ethnicity** | | | |
| Hispanic | 206 (11.6) | 290 (16.8) | 496 (14.2) |
| Non-Hispanic Black | 461 (26.0) | 441 (25.5) | 852 (24.3) |
| Non-Hispanic Other$ | 240 (13.5) | 391 (22.6) | 534 (15.2) |
| Non-Hispanic White | 533 (30.1) | 294 (17.0) | 974 (27.8) |
| Missing | 333 (18.8) | 315 (18.2) | 648 (18.5) |
| **Region** | | | |
| Northeast | 177 (10.0) | 142 (8.2) | 319 (9.1) |
| South | 949 (53.5) | 784 (45.3) | 1733 (49.5) |
| Midwest | 189 (10.7) | 121 (7.0) | 310 (8.9) |
| West | 458 (25.8) | 684 (39.5) | 1142 (32.6) |
| **Diabetes** | 961 (54.2) | 1073 (62.0) | 2034 (58.0) |
| **Vaccination status (number of doses)** | | | |
| 0 | 604 (34.1) | 228 (13.2) | 832 (23.7) |
| 1 | 210 (11.8) | 194 (11.2) | 404 (11.5) |
| 2 | 753 (42.5) | 620 (35.8) | 1373 (39.2) |
| 3 or more | 206 (11.6) | 689 (39.8) | 895 (25.5) |

$Persons self-reporting Asian, American Indian, Alaskan, and Pacific Islander heritages are captured as 'other.'
*IgG index < 10, N=1561; IgG 10-23, N=212

**STable 3. Number of patients included in each 30-day period in Figures 1 A and B.**

| Days since first dose | mRNA1273 | | BNT162b2 | |
|---|---|---|---|---|
| | 1-2 doses | 3+ doses | 1-2 doses | 3+ doses |
| 0-30 | 149 | 1,105 | 669 | 568 |
| 31-60 | 152 | 1,086 | 675 | 560 |
| 61-90 | 142 | 1,128 | 682 | 573 |
| 91-120 | 134 | 1,113 | 681 | 558 |
| 121-150 | 137 | 1,121 | 663 | 549 |
| 151-180 | 134 | 1,121 | 662 | 555 |
| 181-210 | 129 | 1,106 | 669 | 541 |
| 211-240 | 125 | 1,103 | 639 | 525 |
| 241-270 | 107 | 1,041 | 372 | 311 |
| 271-300 | 56 | 694 | 234 | 178 |
| 301-330 | 27 | 336 | 117 | 99 |

**SFigure 1. Boxplots of antibody levels between Feb 1 and Dec 24th 2021 plotted from 1st dose, measured in 30-day periods A. mRNA1273 B. BNT162b2**

**A**

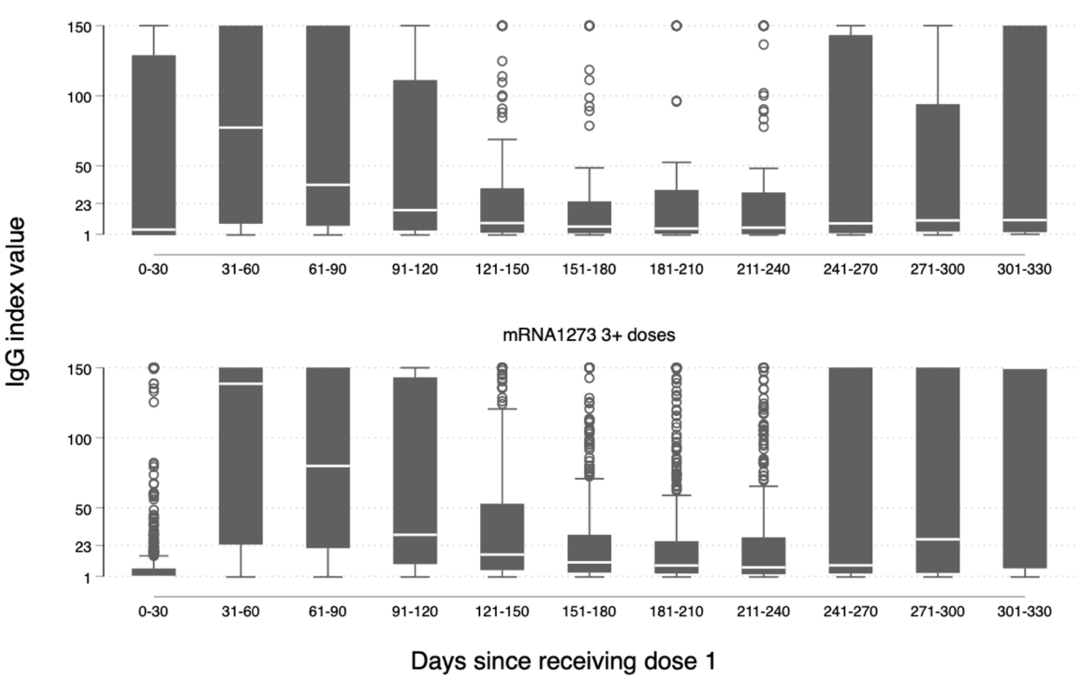


**B**

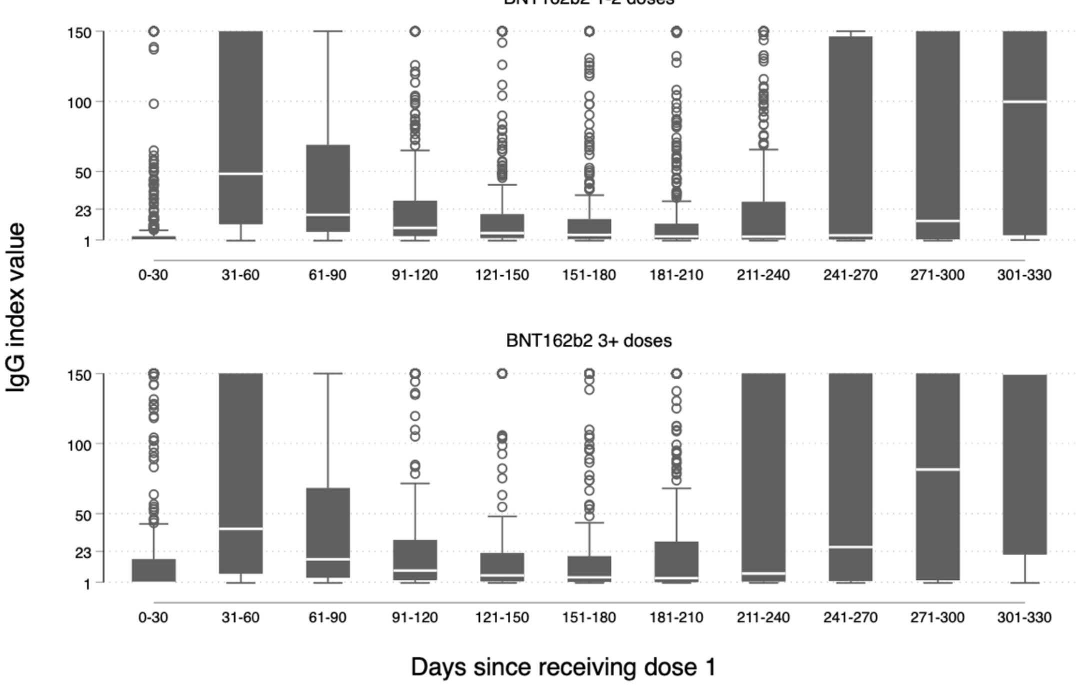
